# An implementation framework to improve the transparency and reproducibility of computational models of infectious diseases

**DOI:** 10.1101/2022.10.28.22281671

**Authors:** Darya Pokutnaya, Bruce Childers, Alice Arcury-Quandt, Harry Hochheiser, Willem G Van Panhuis

## Abstract

Computational models of infectious diseases have become valuable tools for research and the public health response against epidemic threats. The reproducibility of computational models has been limited, undermining the scientific process and possibly trust in modeling results and related response strategies, such as vaccination. We translated published reproducibility guidelines from a wide range of scientific disciplines into an implementation framework for improving reproducibility of infectious disease computational models. The framework comprises twenty-two elements that should be described, grouped into six categories: computational environment, analytical software, model description, model implementation, data, and experimental protocol. The framework can be used by scientific communities to develop actionable tools for sharing computational models in a reproducible way.

## Introduction

Computational models have become valuable tools for the global response against infectious disease outbreaks and pandemics, including the coronavirus disease 2019 (COVID-19) pandemic [1]. Computational models of infectious diseases are representations of biological phenomena in computer code, used to elucidate mechanistic processes of infectious diseases such as transmission and pathogenicity, to study the effect of countermeasures, and to forecast epidemic trajectories [2–4]. As in all scientific research, the validity of a computational model depends on the ability of the community to review and reproduce the modeling “experiment” (i.e., to obtain scientific results consistent with a prior study using the same experimental methods) [5,6].

Reproducibility can be especially limited for computational studies such as computational modeling and artificial intelligence, due to their methodological complexity and heterogeneity of, or restricted access to, data sources [7,8]. The rapid pace of modeling research during the COVID-19 pandemic has raised concerns about the transparency and reproducibility of modeling results [9]. Lack of reproducibility can have serious consequences, as illustrated by retractions of early COVID-19 research from prominent scientific journals and can potentially reduce societal trust in science and consequently, in the public health response against the pandemic [10–12].

Scientific and government agencies, including the US Government Accountability Office and the US National Academies of Sciences, Engineering, and Medicine (NASEM) have published recommendations to enhance the reproducibility of computational research [6,13]. Most recommendations list general principles or suggest types of information that should be reported by scientific publications of computational modeling experiments, such as the checklist recommended by the EPIFORGE guidelines [5,14]. Most guidelines recommend that researchers describe the data sources and modeling methods used and that they share the source code [5,13]. Yet, even with available input data and source code, published results of modeling studies can be difficult or impossible to reproduce [6]. It is notoriously difficult to re-run a published computational model, even when source code is available, because other essential details such as the information about versions and dependencies of the software, or about the required operating system and compute environments, are often missing [15].

Various initiatives have emerged in the scientific community to improve scientific inference based on computational models and to improve public health decision-making during outbreaks. Several initiatives have developed methods for comparing results across multiple models. For example, multi-model comparison studies have been conducted for rotavirus and dengue to better understand the effects of vaccination [16,17]. Recently, multi-model comparison studies for influenza and COVID-19 forecasting have evolved into coordinated projects, such as FluSight and the COVID-19 Forecast Hub, with their own infrastructure for data and model sharing to support decision-making by national and global health agencies (Table S1). New methodology has also been developed to combine results from multiple models into ensemble results to improve epidemic forecasting, e.g., for influenza, Ebola, and COVID-19 [18–20]. In addition, formal decision-analytic methods have been developed to better characterize uncertainty in multi-model comparison projects [21].

The success and impact of model comparison and combination projects depend on methods and technologies that enable researchers to share their modeling experiments in a transparent and reproducible way that characterizes model similarities, discrepancies, and uncertainties. So far, the various model combination projects and modeling hubs have been dependent on ad-hoc methods to describe models and share results. No methodological framework currently exists that can directly be translated into tools for sharing computational models of infectious diseases in a transparent and reproducible manner.

## Results

We developed an implementation framework for representing computational models of infectious diseases in a reproducible format, grounded in previous research on reproducibility from a broad range of scientific disciplines (Fig 1). The framework can be used by researchers and scientific organizations to develop tools and resources, such as checklists and metadata schemas to share transparent and reproducible computational models.

**Fig 1.**
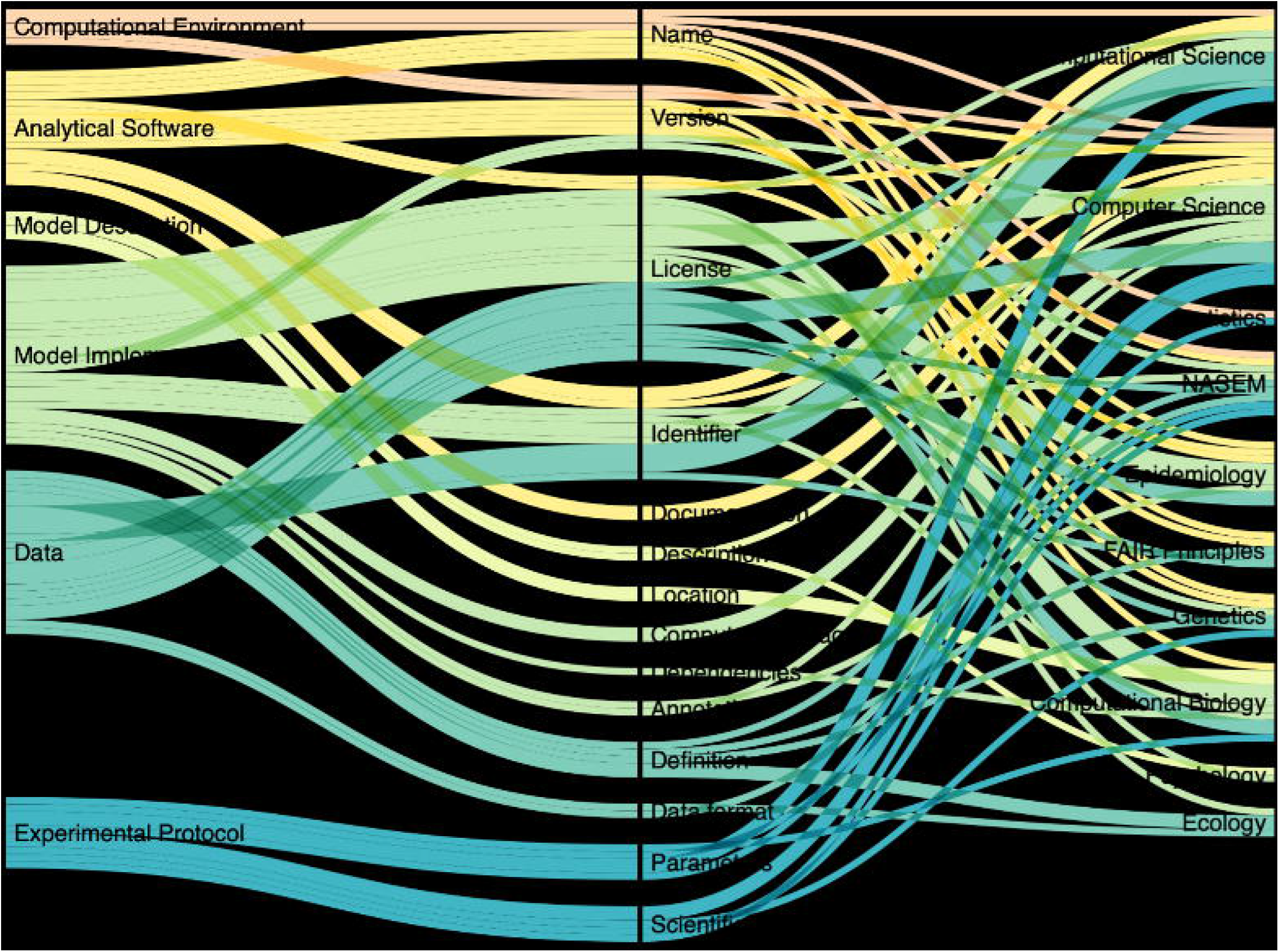
Flow diagram depicting the six implementation framework reproducibility categories (left), their associated elements (center), and the scientific disciplines from which references were identified (right). Width of lines are proportional to the number of references gathered from each scientific discipline to justify inclusion of each category and element.

We identified 22 elements in six categories that together provide a complete representation of the reproducibility for a computational model (Table 1), based on a review of existing guidance on reproducibility and an iterative testing process by our team. The six categories are: (1) computational environment; (2) analytical software; (3) model description; (4) model implementation; (5) data; and (6) experimental protocol. We represented the six categories in a framework that aligns with a commonly used workflow for computational experiments (Fig 2).

**Table 1.** Implementation framework categories, elements, and relevant examples.

| Category and Element | Abbreviated Definition* | Examples |
| --- | --- | --- |
| **1) Computational Environment** |  |  |
| 1.1) Name | Name of operating system | “Microsoft Windows,” “macOS,” “Linux” |
| 1.2) Version | Version of operating system | "Windows 8.1 (Blue)," "macOS 10.12 (Sierra)" |
| **2) Analytical Software** |  |  |
| 2.1) Name | Name of software program | "SAS," "R," "original software name" |
| 2.2) License | Restriction on use | Open source: "R"; propriety: "STATA," "SAS" |
| 2.3) Version | Version of software | "SAS 9.4," "R version 3.3.3" |
| 2.4) Identifier | Unique online identifier | "DOI," "URL" |
| 2.5) Documentation | Availability of documentation to use and install software | URL to installation guide |
| **3) Model Description** |  |  |
| 3.1) Description | Complete, structured description of the model | Equations, diagrams, charts, tables vs. unstructured text |
| 3.2) Location | Specified in the publication/ supplement | Model described in the methods/supplement vs. referenced in preceding publication |
| **4) Model Implementation ("Code")** |  |  |
| 4.1) License | Restriction on use | Publicly stored on GitHub |
| 4.2) Version | Version of code | Version, modification date |
| 4.3) Identifier | Unique online identifier | "DOI," "URL" |
| 4.4) Computer language | Name of code language | "SAS," "R," "STATA," "Python," "C++" |
| 4.5) Dependencies | Additional essential files | Packages, classes, supplementary files |
| 4.6) Annotations | Sufficient, user-interpretable comments | Code suitably annotated for understanding |
| **5) Data** |  |  |
| 5.1) Data calibration | Indicator for whether the model was calibrated to existing vs. simulated data | "Yes" (model calibrated to input data); "no" (model does not require input data) |
| 5.2) Definition | Description of data content and source | Content: column/ field descriptions; source: "CDC," "Project Tycho" |
| 5.3) Identifier | Identifier from where the data are retrieved | "DOI," "URL" |
| 5.4) License | Restriction on use | Publicly stored on GitHub |
| 5.5) Data format | Formatted data for the model implementation | "CSV," "JSON," "XML" |
| **6) Experimental Protocol** |  |  |
| 6.1) Parameters | Description of model parameters | Table or list of parameter values |
| 6.2) Scientific workflow | Process of how categories 1-5 create the results | Explanation in GitHub README.md |

\*Complete definitions provided in the Supplementary Text (S1 Text).

**Fig 2.**
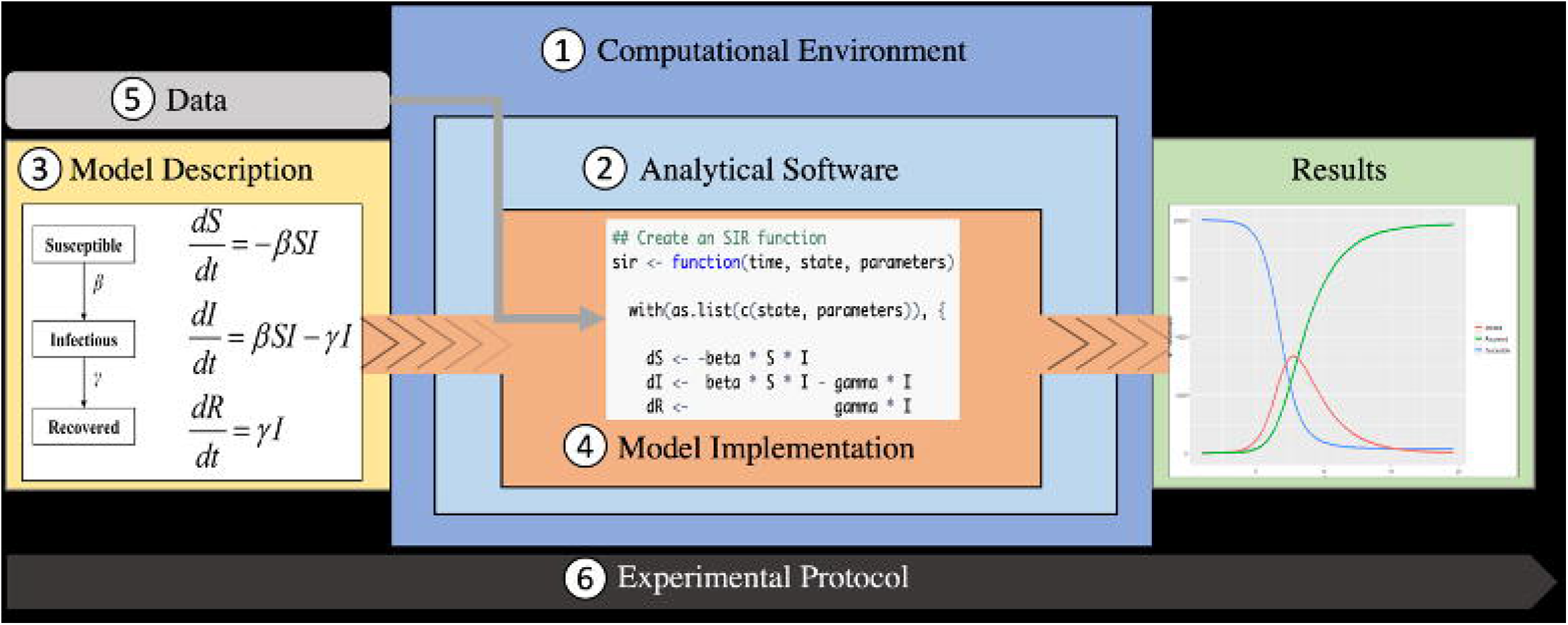
Reproducibility framework. The categories of the reproducibility framework are depicted for the example of a susceptible-infectious-recovered (SIR) model. The SIR model is described using equations in a model description (3) and implemented using a model implementation in the R language (“code”) (4). The code runs in an analytical software, in this case R (2), which runs in some computational environment with an operating system (1). The model implementation can import data (5) and operate on it. The model implementation produces results in the form of new data or visualizations, that leave the analytical software (e.g., PDF) and/or the computational environment (e.g., as printout). The experimental protocol describes the entire workflow and how the categories interact (6).

The computational model is usually described as a combination of narrative text, diagrams, and mathematical equations (model description). The model is then represented in its computational format as a set of commands, functions, and operations encoded in the model source code (model implementation). The data are ingested by the model implementation and operated on to compute the model results. The entire computational experiment is documented in the experimental protocol. A complete description of all the elements will represent a reproducible computational model.

## Discussion

The implementation framework provides a foundation that can be further developed by scientific communities into tools for sharing computational models of infectious diseases in a reproducible way, based on community-specific preferences. For example, the framework can be formalized into a structured metadata schema with prescribed structured vocabularies and ontologies that could render a machine-actionable metadata object compliant with the Findable, Accessible, Interoperable, and Reusable (FAIR) guiding principles [22]. Machine-actionable metadata could enable automated workflows for model comparison and combination efforts and accelerate the use of models for time-sensitive decision making, e.g., in the context of a distributed ecosystem of FAIR data and services, as recently described by Bourne et al. [23]. Beyond just sharing computational models, machine-actionable metadata could become components of dynamic data management and sharing plans (DMSP’s) for computational studies, that could streamline data and model sharing and scientific discoveries [24].

The implementation framework can also be developed into a checklist to represent the degree to which a computational modeling experiment is reproducible, as we did as part of the iterative testing process during the development of the framework (S2 Table). Similar checklists are used to assess the degree of participation of open science at research institutions or the degree of compliance with the FAIR guiding principles [25,26].

Another highly promising method for sharing computational models in a transparent and reproducible manner is through containerization in executable workflow objects, such as Galaxy [27], Open Curation for Computer Architecture Modeling (Occam) [28], and Pegasus [29]. Containerized experiments are especially useful when connected to scholarly publications to ease access, simplify reproduction, and build on prior experiments for new insights. For example, in a pilot project, Occam was connected to the Association for Computing Machinery’s (ACM) Digital Library to demonstrate how containerized workflows can be included and distributed in an executable form with a scholarly article [30]. The implementation framework described in this article specifies what information should be represented in the metadata for containerized workflow objects, so that researchers (and machines) can understand what a certain containerized object represents.

The lack of reproducibility for computational models of infectious diseases can undermine the scientific credibility of modeling results among researchers, policymakers, and even among the public, where skepticism regarding scientific research is already on the rise [31]. It is essential to represent computational models in a transparent and reproducible way so that models can be shared, compared, and combined. We envision that researchers, journals, funders, and scientific organizations can use our framework to develop actionable tools to improve sharing of computational models in a reproducible manner that also accelerates the model-to-decision timeline in response to emerging infectious disease threats.

## Methods

To identify the six categories and 22 elements of the implementation framework, we reviewed guidelines published by the National Academies of Sciences, Engineering, and Medicine (NASEM), the Findability, Accessibility, Interoperability, and Reusability (FAIR) guiding principles, and peer reviewed literature in a variety of scientific domains including general computational science, computer science, statistics, epidemiology, genetics, computational biology, psychology, and ecology (Fig 1).

We identified peer reviewed literature by querying PubMed articles published between January 1st, 2000, and January 1st, 2020, using keywords “reproducible,” “reproducibility,” “computational,” “research,” “data,” and “code.” We limited our search to studies published in English and excluded papers about animal-models or clinical research. We extracted quotes that referenced information that researchers believed was relevant to represent reproducible and transparent computational modeling studies.

We grouped the reproducibility elements into six categories and mapped the relationships between each of the categories to the conceptual model of an infectious disease modeling workflow (Fig 2). The workflow identifies how the six categories are used together to generate the model results. The final implementation framework, along with abbreviated definitions, and relevant examples in infectious disease computational modeling studies are presented in Table 1.

To validate the implementation framework categories and elements, we structured the framework into a checklist (Table S2). The checklist was trialed three times using ten, twenty, and forty-eight infectious disease modeling studies with varying complexities. DP completed the checklist during the first two trials. During the third trial, DP, BC, AAQ, and WP completed the checklist independently. The results of all three trials were reviewed by the entire team. Based on discrepancies between answers, we would modify the checklist to improve clarity and move to the next trial. Iterating through these checklists versions of the implementation framework has improved the real-world application potential of the framework, vs. a more theoretical approach which may not be readily implemented by communities.

For the first trial, ten publications were randomly selected without replacement from all publications authored by Models of Infectious Disease Agent Study (MIDAS) members before September 20, 2019 (n = 3664). Based on title review, if the paper was not related to infectious disease modeling, we randomly selected another paper until we identified ten infectious disease modeling studies. For the second trial, we randomly selected twenty COVID-19 modeling papers without replacement from a list of 229 papers authored by MIDAS members between January 1^st^ - March 31^st^, 2020.

For the third trial, we identified 48 publications. We queried PubMed, medRxiv, bioRxiv,arXiv for COVID-19 modeling publications published between January 1^st^, 2020 – March 31^st^, 2020, using keywords “coronavirus,” “COVID-19,” SARS-Cov-2,” “estimate*,” “model*”, “reproduc*”. The initial query identified 793 records with ten duplicates. The titles and abstracts of the 783 de-duplicated publications were reviewed with inclusion and exclusion criteria. We excluded 224 records based on exclusion criteria: observational, genomic, immunological, and molecular studies, commentaries, reviews, retraction, letter to editor, response articles, not related to COVID-19, clinical trials, and app development. From the remaining 559 papers, we randomly selected 50 publications without replacement for full text review with inclusion and exclusion criteria. A letter to the editor and review paper were excluded. Forty-eight publications were included in the final review process. During the final trial, we randomly assigned team members to each of the 48 publications; each publication was reviewed twice. After the final review, discrepancies between each response were discussed amongst the authors and final edits to the implementation framework and checklist were made.

## Data Availability

All data produced in the present work are contained in the manuscript.

## Acknowledgments

We thank current and past members of the Models for Infectious Disease Agent Study (MIDAS) Coordination Center, including Jessica Kerr, Lucie Contamin, Anne Cross, John Levander, Jeffrey Stazer, Kharlya Carpio, Inngide Osirus, and Lizz Piccoli for critical discussions and feedback. We would also like to thank Tiffany Bogich, a member of the Multi-Model Outbreak Decision Support (MMODS), for her review and feedback during the early development stages of the implementation framework.

## Supporting information

**S1 Text. Supplementary information**. Complete definitions of the implementation framework categories.

**S1 Table.** Examples of multi-model comparison initiatives for influenza and COVID-19 forecasting.

| Collaborative project | Description | URL |
| --- | --- | --- |
| FluSight | Community-based influenza forecasting effort managed by the CDC that combines influenza forecasting models. | https://tinyurl.com/4szpr85y |
| COVID-19 Forecast Hub | Community-based effort for COVID-19 forecasting with a central repository for short-term (1-4 weeks) forecasting data from over 50 research groups. | https://covid19forecasthub.org |
| Multi-Model Outbreak Decision Support (MMODS) | Community-based decision-analytic approach established by the MIDAS community to combine and contrast results from multiple COVID-19 models. | https://midasnetwork.us/mmods/ |
| COVID-19 Scenario Modeling Hub | Modeling hub established to compare and combine models with the objective to project the course of the pandemic in the US several months ahead. | https://tinyurl.com/5n8b4fnf |
| COVID-19 Multi-model Comparison Collaboration (CMCC) | Global effort between modelers and policy makers to compare and interpret COVID-19 models with a focus on models used in developing countries. | https://tinyurl.com/2p9xwk3e |
| US Center for Forecasting and Outbreak Analytics (CFA) | Center established by the CDC to help improve outbreak prediction and response in the US. | https://tinyurl.com/bdzx63ru |
| WHO Modeling Hub in Berlin | Center established by the WHO to help improve outbreak prediction and response globally. | https://tinyurl.com/bdzhfcfw |

Abbreviations: COVID-19, coronavirus disease 2019; CDC, Centers for Disease Control and Prevention; MIDAS, Models for Infectious Disease Agent Study; WHO, World Health Organization

**S2 Table.** Reproducibility framework formatted as a checklist with examples. The checklist consists of questions related to the six categories (1) computational environment; (2) analytical software; (3) model description; (4) model implementation; (5) data; and (6) experimental protocol. The center column provides examples for each category and element.

|  |  |  |
| --- | --- | --- |
| **1. Computational Environment** |  |  |
| 1.1) Is the operating system documented? | Examples: Microsoft Windows, macOS, Linux | ☐ Yes; the operating system is documented<br>☐ No; the operating system is not documented<br>☐ Not applicable |
| 1.2) Is the operating system version documented? | Examples: Windows 8.1 (Blue) or macOS 10.12 (Sierra) | ☐ Yes; the operating system version is documented<br>☐ No; the operating system version is not documented<br>☐ Not applicable |
| **2. Analytical Software** |  |  |
| 2.1) Is the name of the analytical software documented (e.g., the programming language name)? | Examples: SAS, R, STATA, Python, C++. Authors may have also used an originally developed software with a unique name. | ☐ Yes; the name of the analytical software is documented<br>☐ No; the name of the analytical software is not documented<br>☐ Not applicable |
| 2.2) Is the analytical software accessible for free? | For all mentioned analytical software, if the analytical software is available online for download and it is free, mark "Yes." If the analytical software is available online but must be bought or licensed, select "Partially." Additionally, if the authors used multiple types of analytical software in their analyses and not all of them are available for free download, select "Partially." If the analytical software is not available online for download, select "No." Examples of analytical software available for free download: Java, R, Python; non-free examples: STATA, SAS, SPSS, MATLAB. | ☐ Yes; all mentioned software is available for free download<br>☐ Partially; all mentioned software is available for download but requires a purchase and/or acquiring a license or not all the software used were available for free download<br>☐ No; the software is not available for download<br>☐ Not applicable |
| 2.3) Is the version of the analytical software documented? | For all mentioned analytical software, if the version is documented in the publication, supplement, or in an online repository (e.g., GitHub), select "Yes." If the version is documented for some of the mentioned analytical software, select "Partially." If the version is not documented, select "No." Examples: SAS 9.4 or R version 3.3.3. | ☐ Yes; the version is documented for all mentioned analytical software<br>☐ Partially; the version is documented for some mentioned analytical software<br>☐ No; the version of the analytical software is not documented<br>☐ Not applicable |
| 2.4) Do the authors include a specific | If the authors provide a reference for all mentioned software that includes a DOI, | ☐ Yes; all mentioned software is referenced with a specific identifier |
| identifier (DOI, URL, citation) that points to the analytical software that was used? | URL, or version from which the software was retrieved from, select "Yes." If all mentioned software is referenced but the reference does not include the DOI, URL or version, select "Partially." Additionally, if only some mentioned software is referenced, select "Partially." If the authors do not provide a reference for the software, select "No." | such as a DOI, URL, or version<br>☐ Partially; all mentioned software is referenced but the reference does not include the DOI, URL, or version or only some mentioned software is referenced<br>☐ No; none of the mentioned software is referenced<br>☐ Not applicable |
| 2.5) Is the analytical software installation guide accessible online? | For all mentioned analytical software, if the publication includes a link to the installation guide or the installation guide is accessible online, select "Yes." If the installation guide is not accessible online, select "No." Commonly used analytical software such as Java, R, Python, STATA, SAS, SPSS, MATLAB, SAS, STATA have accessible online installation guides. If the authors used an originally developed analytical software, the software may or may not have an accessible installation guide. | ☐ Yes; for all mentioned analytical software, the installation guides are accessible online<br>☐ Partially; for some of the mentioned analytical software, the installation guides are accessible online<br>☐ No; for all mentioned analytical software, the installation guides are not accessible online<br>☐ Not applicable |
| **3. Model Description** |  |  |
| 3.1) Is the complete, structured model description provided in the publication, supplement, or referenced publication? | If there is a complete, structured model description in (e.g., figure, table, or text) in the publication, supplement, or in a referenced publication in a single location, select "Yes." If there are portions of the structured model description throughout the publication, supplement, or referenced publication, but the portions together seem complete, select "Partially." If there is an unstructured but complete model description in the publication, supplement, or referenced publication, select "Partially." If there is an incomplete or no model description, select "No." | ☐ Yes; there is a complete, structured model description in the publication, supplement, or referenced publication<br>☐ Partially; there are portions of the structured model description throughout the publication, supplement, or referenced publication, but the portions together seem complete<br>☐ Partially; there is an unstructured but complete model description<br>☐ No; there is an incomplete or no model description<br>☐ Not applicable |
| 3.2) Is the model specified in the publication or supplement (contrary to being referenced in other papers)? | If the authors used a structured or unstructured model that they described in the publication or its supplemental materials, select "Yes." If the authors cited a previously developed model without describing the model in their publication, select "No." | ☐ Yes; there is a model specified in the publication or supplement<br>☐ No; the authors cited a previously developed model in a referenced paper<br>☐ Not applicable |
| **4. Model Implementation ("Code")** |  |  |
| 4.1) Is the model implementation (e.g., code, workflow) openly accessible online? | If the model implementation (e.g., code, workflow) is provided such that all results (e.g., figures, tables) in the publication can be recreated, select "Yes." A workflow can include instructions of how to | ☐ Yes; the model implementation for all results is openly accessible online<br>☐ Partially; the model implementation for part of the results is openly accessible online |
|  | configure/use a graphical user interface to recreate published results. If the implementation is partially provided, such that only part of the results can be recreated select "Partially." If the model implementation is not provided, select "No." | ☐ No; the model implementation is not openly accessible online<br>☐ Not applicable |
| 4.2) Does the model implementation (e.g., code, workflow) have a version or modification date? | If the version or modification date for the model implementation is documented in the publication, supplement, or in an online repository (e.g., GitHub), select "Yes." If the version or modification date is not provided or it is unclear as to which model implementation the author's used, select, "No." | ☐ Yes; the version or modification date is documented in the publication, supplement, or in an online repository (e.g., GitHub)<br>☐ No; the version or modification date is not provided, or it is unclear as to which model implementation the author's used<br>☐ Not applicable |
| 4.3) Does the model implementation (e.g., code, workflow) have an identifier? | If the model implementation has a unique, persistent, and specific identifier (e.g., DOI or URL to a specific file) or the model implementation is provided in the paper or supplement, select "Yes." If there is a general URL to the model implementation (e.g., a URL to an entire GitHub repository), select "Partially." If the model implementation has no identifier, select, "No." If the model implementation is not available, then select "Not applicable." | ☐ Yes; there is a unique, persistent, and specific identifier (e.g., DOI or specific URL) for the model implementation or the model implementation is provided in the paper or supplement<br>☐ Partially; there is a general URL for the model implementation<br>☐ No; there is no identifier for the model implementation<br>☐ Not applicable |
| 4.4) Is the computer language of the model implementation (e.g., code, workflow) documented? | Often the language of the model implementation will be unambiguous. For example, R uses model implementations written in R language and SAS uses model implementations written in SAS language; however, some publications may use originally developed analytical software. In these situations, it is important for the authors to specify what language(s) the program uses. If the computer language for the model implementation is documented, select "Yes." If the computer language for the model implementation is not documented, select "No." | ☐ Yes; the computer language of the model implementation is documented<br>☐ No; the computer language of the model implementation is not documented<br>☐ Not applicable |
| 4.5) Are all model implementation (e.g., code, workflow) dependencies clearly specified in either the publication or supplemental files? | In some cases, the model implementation may have additional dependencies (e.g., packages or classes). For example, the author's may have imported dplyr or ggplot2 packages in R or the Scanner class in Java in order run the model. Even if the authors do not provide the model implementation, they may still mention the dependencies they used in the methods | ☐ Yes; all the model implementation dependencies are clearly specified in either the publication, supplemental files, or at the top of the model implementation<br>☐ Partially; some of the model implementation dependencies are listed in the publication, supplemental files, or are dispersed throughout the |
|  | section. Analytical software that commonly use packages that are not pre-installed include Java, R, and Python. Analyses in SAS, MATLAB, and STATA occasionally require additional model implementation dependencies. SPSS does not have additional model implementation dependencies. If all the model implementation dependencies are clearly specified in either the publication, supplemental files, or at the top of the model implementation, select "Yes." If some of the model implementation dependencies are listed in the publication, supplemental files, or are dispersed throughout the model implementation, select "Partially." If none of the model implementation dependencies are specified, select "No." | model implementation<br>☐ No; none of the model implementation dependencies are specified<br>☐ Not applicable |
| 4.6) Are the model implementations (e.g., code, workflow) annotated with comments? | If the model implementation is annotated with comprehensible comments that allow the user to easily interpret what the code is doing, select "Yes." If there are no comments accompanying the model implementation, select "No." | ☐ Yes; the model implementations are annotated with comprehensible comments<br>☐ No; the model implementations are not annotated with comments<br>☐ Not applicable |
| **5. Data** |  |  |
| 5.1) Does the model in the publication use input data? | Some models, such as stochastic simulation models, may not require input data. If you answer, "No", you will skip to the next section about the experimental protocol. | ☐ Yes; the model uses input data<br>☐ No; the model does not require input data<br>☐ Not applicable |
| 5.2) Has the content and source of the input data been described in the publication or supplement? | If any of the content (e.g., variable descriptions, number of observations) and source information (e.g., data was from Johns Hopkins, CDC, specific websites, interviews) were described in the publication or supplement for each input dataset, select "Yes." If any of the content and source information was described in the publication or supplement for some input datasets, select "Partially." If any of the content or source information was described for each input dataset, select "Partially." If neither source nor content was described for each input dataset, or only the source or content described for some input datasets, select "No." | ☐ Yes; the source and content have been described for each input dataset<br>☐ Partially; the source and content have been described for some input datasets<br>☐ Partially; the source or content have been described for each input dataset<br>☐ No; neither the source nor content was described for each input dataset, or only the source or content described for some input datasets<br>☐ Not applicable |
| 5.3) Does the paper cite a specific, unique, and | If each input dataset has a unique, persistent, and specific identifier (e.g., a DOI or a URL to a specific file or set of | ☐ Yes; there is a unique, persistent, and specific identifier (e.g., DOI or specific URL to a specific file or set of |
| persistent identifier to refer to each input dataset? | files), select "Yes." A specific URL would point to a data file, a version, and/or specific download date to enable re-downloading of the same data used in the analysis. Additionally, if the data are provided in the publication or supplement itself, select "Yes." If there is a general URL to each input dataset (e.g., a URL to a GitHub repository but that it is unclear as to which files are used in the analysis or a web-based data archive), select "Partially." If only some of the input datasets have a unique, persistent, and specific identifier or a general URL, select "Partially." If none of the data described have an identifier, select, "No." | files), or the data are included in the paper or supplement<br>☐ Partially; there is a general URL (e.g., a URL to a GitHub repository or online data archive) for data or used in the analysis<br>☐ Partially; some of the input data have unique, persistent, and specific identifiers<br>☐ No; none of the input data have an identifier<br>☐ Not applicable |
| 5.4) Is the input data openly accessible? | If all the mentioned data used to create the results (e.g., figures and/ or tables) are openly accessible in either the publication, supplement, a specified online repository (e.g., GitHub), or can be downloaded from a specified website (e.g., Johns Hopkins dashboard, CORD-19, Project Tycho), select "Yes." If some of the mentioned data used to create the results are provided in either the publication, supplement, online repository, or can be downloaded from a website, select "Partially." If the data are not openly accessible, or if the authors state that the data are not available, select, "No." | ☐ Yes; all the mentioned data used in the analysis are openly accessible in either the publication, supplement, online repository, or can be downloaded from a website<br>☐ Partially; part of the mentioned data used in the analysis are openly accessible in either the publication, supplement, online repository, or can be downloaded from a website<br>☐ No; none of the mentioned data used to create the results are openly accessible<br>☐ Not applicable |
| 5.5) Is the data in a format that can be easily re-formatted (or "parseable") to meet the input specifications of the model implementation? | If all the mentioned input data are in a format that can be easily re-formatted (e.g., CSV, JSON, XML, PDF table that can be copied as a CSV), select "Yes." If some of the mentioned input data are in a format that can be easily re-formatted, select "Partially." If the data are in a format that is not easily re-formatted (e.g., narrative text, PDF image file, PNG), select "No." | ☐ Yes; all the input data are in a format that can be easily re-formatted<br>☐ Partially; some of the input data are in a format that can be easily re-formatted<br>☐ No; all data are in a format that is not easily re-formatted<br>☐ Not applicable |
| **6. Experimental Protocol** |  |  |
| 6.1) Are all the mentioned parameter values for the model implementation (e.g., code, workflow) documented in a | If the authors provide the parameter values for the model implementation in a single location (e.g., table or list) in the text or supplement, select "Yes". If the authors provide some of the parameter values or the parameter values are dispersed throughout the text or supplement, select "Partially". If the authors do not provide | ☐ Yes; all mentioned parameter values for the model implementation are documented in a single location<br>☐ Partially; some mentioned parameters for the model implementation are documented or they are dispersed throughout the text<br>☐ No; none of the parameter values |
| single location (e.g. table or list in the publication or supplement)? | any of the parameter values, select "No". | for the model implementation are documented<br>☐ Not applicable |
| 6.2) Is there an explanation of how the described/mentioned components (computational environment, analytical software, model implementation, and data) were used together to create the results (e.g., figures and/ or tables)? | If there is a complete explanation in the publication, supplement, or online repository for how all the components (computational environment, analytical software, model description, model implementation, data) were used to create the results, select "Yes." If there is a partial explanation in the publication, supplement, or online repository for how all the mentioned were used to create the results, select "Partially." If there is no explanation of how the mentioned components were used together to create the results, select "No." If the authors did not mention more than one of the previous components, select "Not applicable. | ☐ Yes; there is complete explanation in the publication, supplement, or online repository for how all the mentioned components were used to create the results<br>☐ Partially; there is a partial explanation or how the data and model implementation were used to create the key results<br>☐ No; there is not an explanation of how the data and model implementation were used to create the key results<br>☐ Not applicable |

