## Supplementary Text for "An implementation framework to improve the transparency and reproducibility of computational models of infectious diseases"

### Complete definitions of the implementation framework categories

The final reproducibility framework consisted of six categories and their associated 22 elements that each need to be described to represent a reproducible computational model: (1) computational environment; (2) analytical software; (3) model description; (4) model implementation; (5) data; and (6) experimental protocol.

#### 1. Computational environment

The computational environment comprises the software, hardware, hardware drivers, operating system, libraries, and environmental variables that are required to conduct the study (*32*). Failure to reproduce modeling studies, even if the data and code have been made available, can be due to incompatibilities or specific requirements in the computational environment. We emphasized documenting the operating system (e.g., MacOS, Linux, Microsoft Windows) given that certain analytical software require a specific operating system to run (e.g., SAS software requires Microsoft Windows).

#### 2. Analytical software

Analytical software is the software program used to implement the model, such as Java, R, Python, and MATLAB. Scientists may use either custom or previously developed software that is either open-source or propriety. Studies using propriety software can be reproduced but these reproductions use more resources (effort and financial) compared to similar efforts that use open-source software (*33*). In addition to the name and version of the analytical software, a specific identifier such as a DOI, URL, or a citation from where the software was retrieved should be provided in the publication or supplement, to allow others to access the same version of the software and to find the software documentation, including the installation guide. In some cases, the model implementations have additional software dependencies such as software programs, libraries, specific packages, classes, or modules. Ideally, a table or list of dependencies would be provided either in the supplement or at the top of the model implementation itself.

### 3. Model Description

Authors describe their models either in a structured (equations, diagrams, charts, tables) or unstructured (text) format, or both, and sometimes refer to previously published model descriptions. Ideally, authors would provide a structured diagram or complete description of the model. For example, for a compartment model, the diagram and description would represent the states and transitions between compartments, along with the corresponding equations. A structured model description in a single location (vs. distributed throughout a paper) would be the easiest to understand, reproduce, and be least prone to misinterpretation.

### 4. Model Implementation ("Code")

Model implementations are a set of executable commands written in a programming language to process and analyze data. The term "model implementation" may also be referred to as the "code." Here, we chose the term "model implementation" to distinguish the code written by the user to implement their model in an analytical software from other types of code, such as the "code" used by the operating system and other parts of the computational environment. Openly accessible, versioned model implementations should be documented in an online repository (e.g.,

GitHub) that is specifically referred to in the publication and/or supplement. Model implementations should be accompanied by comprehensible annotations that allow the user to interpret the purpose of the model implementations.

### 5. Data

To reproduce the key (published) results of a modeling study, the authors may have fit the model to existing, original, or simulated data. In some cases, models are based on assumptions only and are not fit to any data. If data were used, their source and content, including descriptions of the columns or fields, should be described in the publication or supplement. Datasets should be available in such a way that others can derive the exact data used for the study, in the format required by the input specification of the model implementation (e.g., CSV, JSON, XML). Data do not have to be openly accessible to achieve reproducibility; however, open data are easier to reproduce compared to studies which require researchers to take additional steps (e.g., licensure) to acquire the data. Ideally openly accessible data would have their own registered unique, persistent identifier to prevent any breaks or changes in linked URLs (e.g., DOI).

### 6. Experimental Protocol

The experimental protocol is the description of the workflow explaining how all the previously listed categories and the model parameter values are used together to create the (published) results (e.g., a well-documented README.md file in GitHub). In simple analyses, the experimental protocol might be obvious; however, in complex studies with multiple software and datafiles, a clear experimental protocol can save researchers time during future attempts to reproduce a study. As part of the experimental protocol, model parameters should be placed in a single location such as a table or list in the publication or supplement. For example, for compartmental models, the effective contact rate (β) and the rate of recovery (γ) are two parameters that should be clearly specified.
